# Contribution of mitochondrial DNA heteroplasmy to the phenotypic variability in maternally transmitted 22q11.2 deletion syndrome

**DOI:** 10.1101/2020.05.31.20118646

**Authors:** Boris Rebolledo-Jaramillo, Maria Gabriela Obregon, Victoria Huckstadt, Abel Gomez, Gabriela M. Repetto

**Affiliations:** Center for Genetics and Genomics, Facultad de Medicina Clínica Alemana, Universidad del Desarrollo, Santiago, Chile.; Servicio de Genética, Hospital de Pediatría Garrahan, Buenos Aires, Argentina.

## Abstract

22q11.2 deletion syndrome (22q11DS) has an incidence of 1 in 4,000. Most cases occur de novo, but about 10–15% of cases are inherited. Features include congenital heart disease, cleft palate, developmental delay, and other characteristics that can vary even among family members. The presence of nuclear mitochondrial genes in the deleted region, and the requirement of mitochondrial function for proper embryonic development, suggests that intrafamilial variability in maternally transmitted 22q11DS could be explained, at least partially, by variation in mitochondrial DNA (mtDNA). Thus, we sequenced the mtDNA of seventeen 22q11DS mother-child pairs. We identified 29 heteroplasmic variants at the 1% level, and compared the intrafamilial allele frequency change between phenotypically concordant and discordant pairs. We observed a statistically significant difference for the palatal phenotype: p-value = 0.048 (permutation test, 8 concordant vs. 9 discordant pairs), but not for the cardiac phenotype: p-value = 0.568 (6 vs. 11).

Mitochondrial function has been primarily studied in mouse models of neurological 22q11DS phenotypes. Our study sets a precedent for considering human mitochondrial variation as a genetic modifier of congenital defects in this syndrome, and although our results are limited by sample size, they suggest a role for mitochondrial variation in the palatal phenotype.

## INTRODUCTION

Chromosome 22q11 microdeletion syndrome (22q11DS) is one of the most frequent pathogenic genomic rearrangements in humans with an estimated incidence of 1 in 4,000 live births^1^. Clinical features include congenital heart disease in 50–70% of patients, cleft palate in 70–80%, developmental delay and learning difficulties in virtually all patients, increased risk of psychiatric disease, relative short stature and recognizable facial features, among many other characteristics^2^. Through knock-out experiments in mice, it was shown that haploinsufficiency for the transcription factor Tbx1, one of the genes located within the deletion region, reproduces the cardiac outflow tract anomalies, cleft palate and thymic and parathyroid hypoplasia, similar to the clinical phenotype observed in 22q11DS patients^3^. The syndrome is characterized by the variable expressivity of its phenotypes, illustrated by descriptions of phenotypic “extremes” that range from very ill newborns manifesting severe conotruncal heart disease, hypocalcemic seizures, cleft palate and immunodeficiency, to individuals with a history of learning disabilities and hypernasal speech due to a submucous cleft palate, or adolescents or adults with schizophrenia and no structural anomalies. Most individuals share a deletion of the same size, however deletion size does not appear to be associated with specific manifestations^2,4^. The cause of this variability is still unknown, and the causes may be genetic (inherited or acquired), epigenetic, environmental or stochastic. Interestingly, a “two-hit hypothesis” has been proposed as a mechanism for variable expressivity for other microdeletion syndromes, where the severity of the phenotype of 15q13 and 16p12.1 microdeletions appears to be increased in the presence of a second genetic or genomic alteration^5^. Therefore, in the search for potential second hits, some researchers have focused on characterizing the pathways where TBX1 is involved. Using proteomics, it was shown that TBX1 upregulates expression of proteins primarily involved in metabolism, and downregulates expression of proteins involved in signaling, and interestingly, between 10–15% of these identified proteins localized to the mitochondria^6^. There are six nuclear mitochondrial genes within the boundaries of the common 3-Mb 22q11.2 deletion^1^. As described in mice, multiple energy-demanding stages of cell migration occur between E6.5 and 9.5 to form the heart^7^. In mice knockout for Polg2, the mitochondrial DNA (mtDNA) polymerase, development stops at E8.5, around the time the linear heart undergoes looping^7,8^, suggesting that proper mitochondrial function is critical for embryonic development. Similarly, it has been proposed that impaired mitochondrial function contributes to the initial establishment of the 22q11DS neurological phenotype early during development^9^.

In humans, the mtDNA is inherited from the mother alone. mtDNA mutations usually affect only a proportion of the total mitochondrial genomes, generating a state called heteroplasmy^10^. It has been observed that as the percentage of mtDNA harboring a pathogenic variant increases, mitochondrial function declines, and when energy output is insufficient, a threshold is crossed and symptoms appear^11,12^. However, because of their non mendelian inheritance, if mutated mtDNA genomes are present in a mother, the level of heteroplasmy inherited by her children can vary from 0% to 100% in a single generation, due to a phenomenon called the “germline bottleneck”^13–15^, making it difficult to anticipate their effect.

The 22q deletion usually occurs de novo, but it can be also inherited from an affected parent in about 10% of the cases. Similar to unrelated cases, there is intrafamilial variation, with some members of the same family showing more severe phenotypes than others^16,17^, including phenotypically discordant twins^18^. In cases of maternal inheritance of the 22q11 deletion, both the deleted region and the mitochondrial genome were inherited from the mother, yet in some cases only the child presents palate anomalies or cardiac features associated with the syndrome^19,20^. The presence of nuclear mitochondrial genes in the deleted region, and the requirement of mitochondrial function for proper embryonic development, suggest that the observed intrafamilial variability in the 22q11 syndrome could be explained, at least in part, by variation in the mitochondrial DNA. Thus, we explored the extent of mitochondrial DNA variability among del22q11 mother-child pairs discordant for congenital heart disease and palate anomalies.

## METHODS

### Subjects

Mother-child pairs with FISH or MLPA confirmed cases of 22q11.2 deletion, and available information on their cardiac and palate phenotype, were invited to participate. Informed consent was obtained for all participants. This study was conducted under an Internal Review Board-approved protocol at the Universidad del Desarrollo, Chile and Hospital Garrahan, Argentina.

Mothers and their children were initially evaluated by a clinical geneticist at Hospital Garrahan (GH). Children suspect of cardiac or palate anomalies were further evaluated by a cardiologist or an ENT doctor, accordingly. The cardiac and palate phenotype of mothers were obtained from their self-reported medical history.

Sample phenotype description is shown in Table 1. Pairs were considered phenotypically “concordant” when both members shared the presence or absence of any cardiac or palatal structural anomalies, and “discordant”, when one of the two had anomalies and the other member did not.

**Table 1.** Clinical phenotype of patients included in this study. ASD: Atrial septal defect. BU: Bifid uvula. CP: Cleft palate. IIA: Interrupted aortic arch. MVP: Mitral valve prolapse. SMCP: Submucous cleft palate. TOF: Tetralogy of Fallot. TA: Truncus arteriosus. VPI: Velopharyngeal insufficiency. VSD: Ventricular septal defect.

| Pair | Sample | Relationship | Palate | Cardiac |
| --- | --- | --- | --- | --- |
| 1 | DG18 | child | VPI/BU | TA |
|  | DG213 | mother | normal | MVP |
| 2 | DG33 | child | VPI | normal |
|  | DG34 | mother | VP | normal |
| 3 | DG250 | child | VPI | IIA |
|  | DG60 | mother | VPI/BU | normal |
| 4 | DG233 | child | VPI | ASD |
|  | DG76 | mother | normal | normal |
| 5 | DG132 | child | VPI | VSD |
|  | DG133 | mother | normal | normal |
| 6 | DG139 | child | VPI | TOF |
|  | DG161 | mother | normal | normal |
| 7 | DG179 | child | normal | TA |
|  | DG180 | mother | normal | normal |
| 8 | DG185 | child | VPI | VSD |
|  | DG184 | mother | normal | normal |
| 9 | DG193 | child | VPI | ASD |
|  | DG184 | mother | normal | normal |
| 10 | DG221 | child | VPI | normal |
|  | DG220 | mother | normal | normal |
| 11 | DG222 | child | VPI | normal |
|  | DG220 | mother | normal | normal |
| 12 | DG224 | child | normal | normal |
|  | DG225 | mother | normal | normal |
| 13 | DG231 | child | SMCP | TOF |
|  | DG232 | mother | VPI | Murmur |
| 14 | DG236 | child | normal | TA |
|  | DG237 | mother | normal | VSD |
| 15 | DG242 | child | VPI | TA |
|  | DG243 | mother | VPI | normal |
| 16 | DG246 | child | VPI | VSD |
|  | DG247 | mother | CP | normal |
| 17 | DG249 | child | normal | TA |
|  | DG248 | mother | CP | normal |

### DNA extraction and sequencing

Total DNA from blood was submitted for 2×100 DNBSeq^™^ Small genome sequencing at BGI Americas Corporation. According to BGI’s protocol, mitochondrial DNA was enriched using a long-range PCR reaction with primers: forward (MT:15149–15174) TGAGGCCAAATATCATTCTGAGGGGC, and reverse (MT:14816–14841) TTTCATCATGCGGAGATGTTGGATGG, Phusion High Fidelity DNA Polymerase and 5x Phusion GC buffer (New England Biolabs).

### mtDNA variant calling

The heteroplasmy variant calling pipeline has been previously published^14^. In short, raw sequencing reads were aligned with bwa (Li & Durbin, 2009) against the hg19 version of the human genome, but replacing the mitochondrial genome with the revised Cambridge Reference Sequence (rCRS, NC_012920). Alignments were refined using Samtools (Li et al., 2009) for manipulation, Picard Tools for deduplication, GATK for base quality score recalibration^23^, and bamleftalign, from the Freebayes package, for indel standardization^24^. Improved bam formatted files were processed with a custom script that utilizes the Naïve Variant Caller and Allele Counts tools^25^, to extract per nucleotide allele counts for all 16,569 human mtDNA positions. High quality heteroplasmic sites were defined as those sites with: (1) depth ≥ 1,000, (2) minor allele frequency ≥ 1%, (3) no strand bias, (4) outside known problematic regions (303–311, 3107, 16185–16193), (5) no position-in-read bias, and (6) statistically significant in a Poisson test comparing allele frequency to error rate at the position among all the other samples.

### mtDNA haplogroup assignment

We extracted the most common allele for each position in the mtDNA of a sample based on the count of A’s, C’s, G’s and T’s at a position. Then, we concatenated all alleles into a single string of 16,569 letters to create a major-allele sequence for each sample in fasta format. Haplogroup assignment was then calculated on a sample’s major-allele fasta file using the web version of Haplogrep at https://haplogrep.uibk.ac.at^26^.

### Comparison of concordant and discordant mother-child pairs

We counted the number of heteroplasmic sites observed in a mother and her child, calculated the difference between them, and compared concordant and discordant pairs. Zero indicated no change in the number of sites, negative values indicated more sites in the mother, and positive numbers indicated that children had more sites than their mothers.

We tabulated each variable site in pairs (supplementary table 2) and extracted the allele frequency for the one allele identified as heteroplasmic. Then, we calculated the allele frequency change (∆AF) between a mother and her child as ∆AF = AF_child_ – AF_mother_, and compared the distribution of ∆AF values among concordant and discordant pairs. Positive values indicate greater allele frequency in the child than in the mother.

### Statistical analyses and results reproducibility

Descriptive statistics are presented and indicated as mean ± s.d. or median and range. The Shapiro-Wilk test was used to asses normality. The p-value for the comparison of concordant and discordant pairs was calculated with a 1,000 replicas permutation test of the Mann-Whitney U statistic. Results were considered statistically significant if p ≤ 0.05. A Jupyter notebook^27,28^ with the code demonstrating all the steps in data processing, statistical analysis, and figure generation is available at https://github.com/berebolledo/mtDNA22q.

## RESULTS

### Subjects

Thirty-two patients participated in the study, 15 mothers and 17 children, corresponding to 17 mother-offspring pairs with the 22q11.2 microdeletion. Two mothers (DG184 and DG220) had two children each with the deletion. The median and age range of the mothers was 29 [19 – 42] years, and 6 months [1 month – 9 years] for the children. Among the children there were 9 girls (53%), and 8 boys (47%). Two mothers (13.3%) had a CHD, while this feature was present in 13/17 (76.5%) of the children. Six mothers (40%), and 13/17 (76.5%) children had a diagnosis of palate anomalies. For the cardiac phenotype, there were 6 concordant mother-child pairs, and 11 discordant pairs. Based on the palatal phenotype, there were 8 concordant mother-child pairs, and 9 discordant pairs. A summary of palatal and cardiac manifestations is presented in Table 1.

### Sequencing results

On average, 16,322 ± 89 mtDNA positions were captured per sample (mean ± sd), with a median depth of 47,000x (Supplementary figure 1). We identified a particular drop in sequencing depth across all samples around position MT:15,000 consistent with the location of the primers used for mtDNA enrichment (supplementary figure 2, Methods). Therefore, we excluded the region MT:14800–15200 from the remaining analyses.

### mtDNA variants in mother/child pairs

Mitochondrial DNA haplogroups were determined with Haplogrep (Weissensteiner et al., 2016, Methods). The average posterior probability of haplogroup assignment was 0.9 ± 0.07 (mean ± sd). We confirmed all mother-child pair maternal relationships based on their haplogroup (supplementary table 1), and their mtDNA sequence pairwise distance in a neighbor joining tree (supplementary figure 3).

Heteroplasmic variants were calculated as described before in Rebolledo-Jaramillo (2014). We found 29 high confidence sites with minor allele frequency (MAF) ≥ 1% among 32 samples (median number of sites per sample 1, range 0–5). The unusual minor allele frequency distribution of samples DG224 and DG33 (supplementary figure 4), both with five sites each, raised the suspicion of possible sample contamination. Consequently, we evaluated this possibility using a phylogenetic approach^29^, and found found no evidence of sample cross-contamination for neither sample (supplementary figure 5).

These 29 sites were distributed as follows: 5 in the D-loop, 20 in protein coding genes, 2 in RNA genes, and 2 in intergenic regions (table 2). Only one of the sites was considered “confirmed pathogenic” according to MITOMAP^30^: change G12315A found in sample DG224 is associated with atherosclerosis^31^.

**Table 2.**
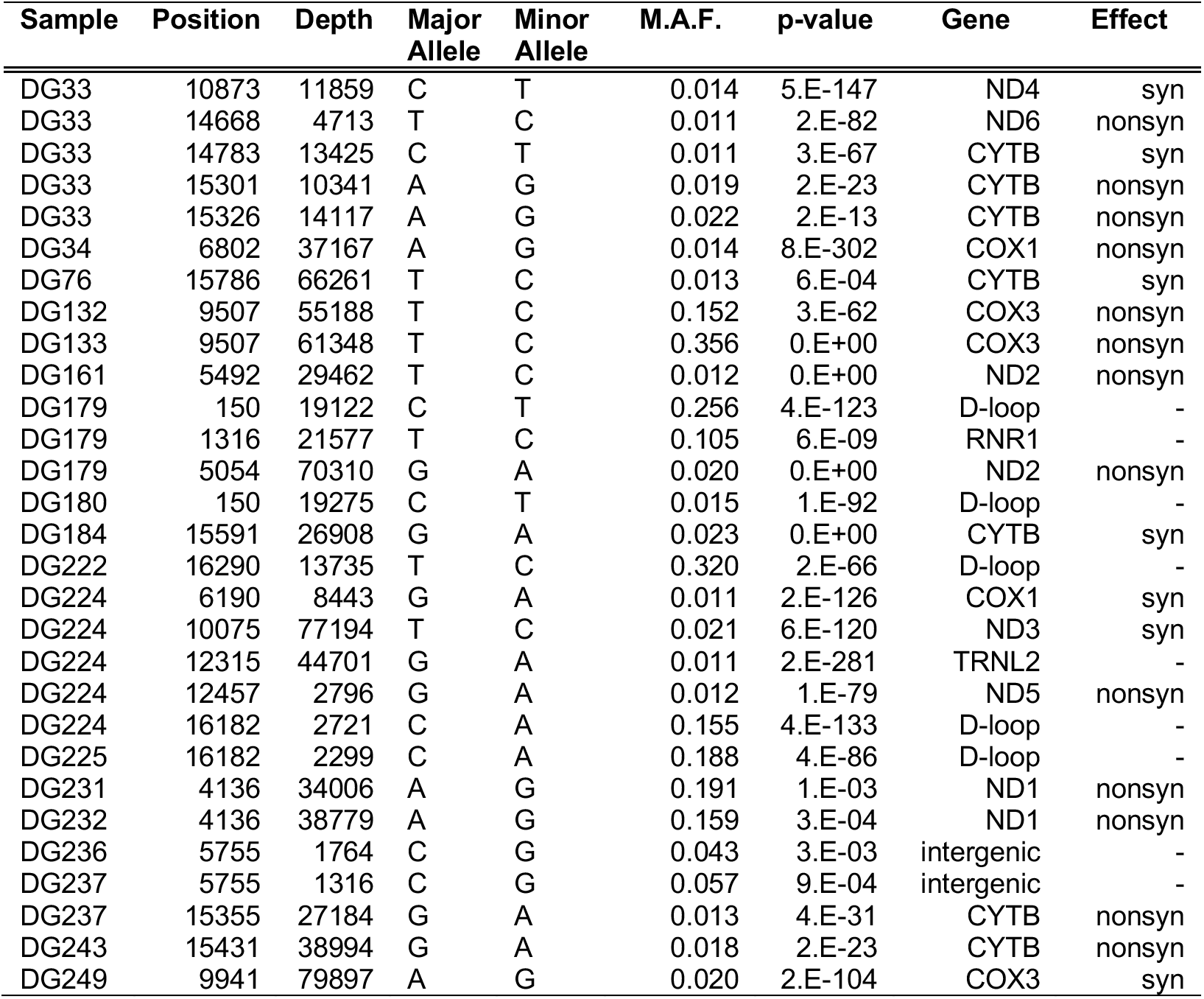
High quality heteroplasmic sites. M.A.F: minor allele frequency. P-value: Poisson test comparing error rate and frequency.

| Sample | Position | Depth | Major Allele | Minor Allele | M.A.F. | p-value | Gene | Effect |
| --- | --- | --- | --- | --- | --- | --- | --- | --- |
| DG33 | 10873 | 11859 | C | T | 0.014 | 5.E-147 | ND4 | syn |
| DG33 | 14668 | 4713 | T | C | 0.011 | 2.E-82 | ND6 | nonsyn |
| DG33 | 14783 | 13425 | C | T | 0.011 | 3.E-67 | CYTB | syn |
| DG33 | 15301 | 10341 | A | G | 0.019 | 2.E-23 | CYTB | nonsyn |
| DG33 | 15326 | 14117 | A | G | 0.022 | 2.E-13 | CYTB | nonsyn |
| DG34 | 6802 | 37167 | A | G | 0.014 | 8.E-302 | COX1 | nonsyn |
| DG76 | 15786 | 66261 | T | C | 0.013 | 6.E-04 | CYTB | syn |
| DG132 | 9507 | 55188 | T | C | 0.152 | 3.E-62 | COX3 | nonsyn |
| DG133 | 9507 | 61348 | T | C | 0.356 | 0.E+00 | COX3 | nonsyn |
| DG161 | 5492 | 29462 | T | C | 0.012 | 0.E+00 | ND2 | nonsyn |
| DG179 | 150 | 19122 | C | T | 0.256 | 4.E-123 | D-loop | - |
| DG179 | 1316 | 21577 | T | C | 0.105 | 6.E-09 | RNR1 | - |
| DG179 | 5054 | 70310 | G | A | 0.020 | 0.E+00 | ND2 | nonsyn |
| DG180 | 150 | 19275 | C | T | 0.015 | 1.E-92 | D-loop | - |
| DG184 | 15591 | 26908 | G | A | 0.023 | 0.E+00 | CYTB | syn |
| DG222 | 16290 | 13735 | T | C | 0.320 | 2.E-66 | D-loop | - |
| DG224 | 6190 | 8443 | G | A | 0.011 | 2.E-126 | COX1 | syn |
| DG224 | 10075 | 77194 | T | C | 0.021 | 6.E-120 | ND3 | syn |
| DG224 | 12315 | 44701 | G | A | 0.011 | 2.E-281 | TRNL2 | - |
| DG224 | 12457 | 2796 | G | A | 0.012 | 1.E-79 | ND5 | nonsyn |
| DG224 | 16182 | 2721 | C | A | 0.155 | 4.E-133 | D-loop | - |
| DG225 | 16182 | 2299 | C | A | 0.188 | 4.E-86 | D-loop | - |
| DG231 | 4136 | 34006 | A | G | 0.191 | 1.E-03 | ND1 | nonsyn |
| DG232 | 4136 | 38779 | A | G | 0.159 | 3.E-04 | ND1 | nonsyn |
| DG236 | 5755 | 1764 | C | G | 0.043 | 3.E-03 | intergenic | - |
| DG237 | 5755 | 1316 | C | G | 0.057 | 9.E-04 | intergenic | - |
| DG237 | 15355 | 27184 | G | A | 0.013 | 4.E-31 | CYTB | nonsyn |
| DG243 | 15431 | 38994 | G | A | 0.018 | 2.E-23 | CYTB | nonsyn |
| DG249 | 9941 | 79897 | A | G | 0.020 | 2.E-104 | COX3 | syn |

### Comparison of concordant and discordant pairs

We hypothesized that, since children inherit both the 22q11.2 deletion and the mitochondrial genome from their mother, changes in the number and/or allele frequency of mtDNA variants could be associated with their phenotype discordance. The difference in the number of sites in cardiac concordant and discordant pairs was 0.5 [-1, 4], and 0 [-1, 2], respectively (median and range). Similarly, the difference in palate concordant and discordant pairs was 0 [-1, 4], and 0 [-1, 1]. Neither comparison resulted statistically significant: p-value 0.058 for the cardiac, and 0.152 for the palate phenotypes, respectively. Next we calculated the allele frequency change (∆AF) between a mother and her child for all heteroplasmic sites identified (Methods and supplementary table 2), and compared concordant and discordant pairs. For the cardiac phenotype, concordant pairs had ∆AF = 0.01 [-0.60, 0.09] (median and range), and discordant pairs had ∆AF = –0.012 [-0.20, 0.24], p-value = 0.568. For the palate phenotype, concordant pairs had ∆AF = 0.01 [-0.03, 024] (median and range), and discordant pairs had ∆AF = –0.02 [-0.60, 0.09], p-value = 0.048 (Figure 1).

**Figure 1.**
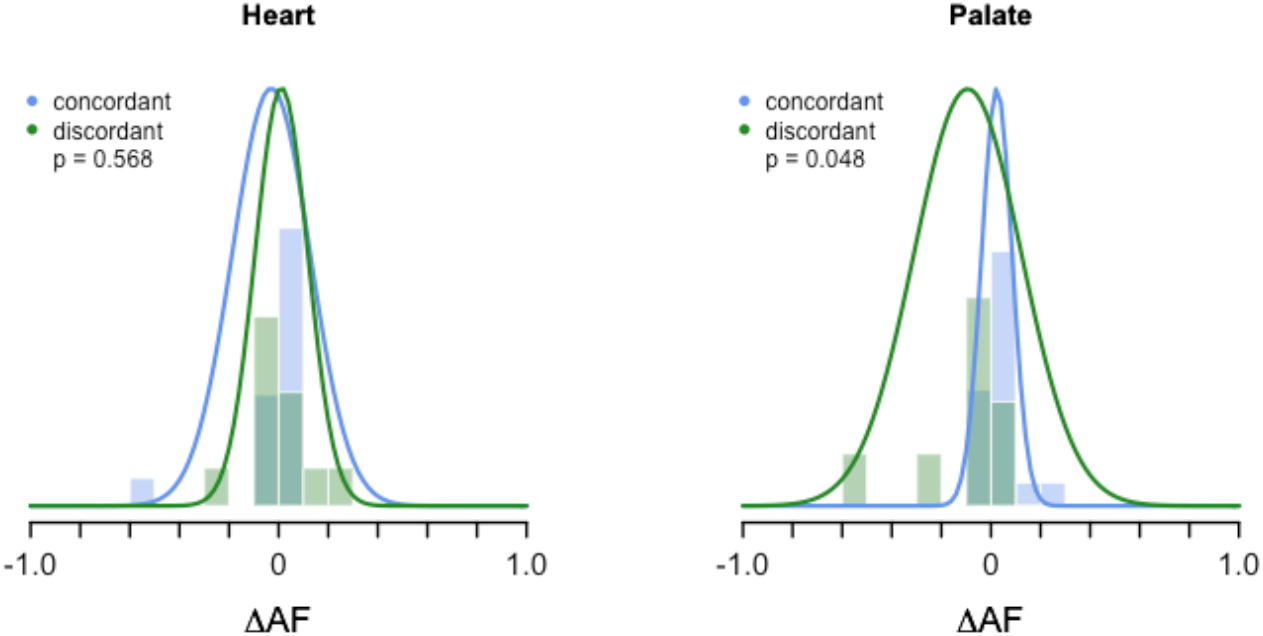
Heteroplasmic allele frequency change (∆AF) between a mother and her child. The histograms describe the actual distribution of the data, whereas the curves represent the expected density distribution provided each sample’s mean and standard deviation.

## DISCUSSION

This study analyzed the contribution of mitochondrial DNA variability to the intrafamilial variation in cardiac and palatal anomalies observed in 22q11.2 deletion patients. We found no differences between phenotypically concordant and discordant pairs in terms of the number of mtDNA variants observed in a mother and her child, or the allele frequency change (∆AF) for the cardiac phenotype, but we did observe a marginally significant difference in ∆AF between concordant and discordant pairs affected by palatal anomalies.

Evaluating the contribution of mitochondrial DNA variants to the cardiac or palatal phenotypes is novel, since mitochondrial dysfunction has been primarily studied in the context of the neurological phenotypes associated with the 22q11DS (Devaraju & Zakharenko, 2017; Li et al., 2019; Napoli et al., 2015), disregarding the mitochondria’s pivotal role during the development of multiple other structures, including the heart^7,8^.

The frequency of congenital cardiac and palatal anomalies in our patients reflected the frequency observed in larger cohorts^1^. And as expected, mothers had lower frequencies of anomalies compared to their offspring, possibly due to cohort effects, and the lower survival and reproductive fitness observed in individuals with heart defects^35^. Our results are limited by the small sample size, as most studies of rare disorders are, and thus underpowered to detect small effects. Nevertheless, the distribution of ∆AF observed in palatal concordant and discordant pairs is consistent with the hypothesis that mtDNA variation might contribute to the phenotypic variability observed in 22qDS patients. These results, however, do not imply causality since most of the variants observed here have not been previously described as pathogenic in MITOMAP^30^. Pathogenicity of mitochondrial variants and heteroplasmic levels are hard to determine, especially in the era of high throughput sequencing where the detection of low level co-occurring mitochondrial variants is common^36^. Moving forward, since these preliminary results suggests a connection between non-pathogenic heteroplasmic levels differences and the palatal phenotype, it could be promising to analyze the impact of non-pathogenic mtDNA in clinical phenotypes. In fact, it was recently reported that a particular haplotype of non-pathogenic mitochondrial missense variants segregated in two families with Leber’s hereditary optic neuropathy (LHON), suggesting that not necessarily a single pathogenic variant, but multiple missense variants can lead to reduced mitochondrial function and trigger a clinical phenotype^37^.

Another source of mitochondrial variation that would be worth exploring in the context of complex phenotypes is the so called “mito-nuclear discordance” (MND). MND refers to the difference in the ancestral origin of the mtDNA and nuclear mitochondrial genes, and it is known to cause changes in OXPHOS efficiency in model organisms^38^. However, whether these incompatibilities naturally exist and contribute to phenotypic variation in humans is currently unknown, but some signals of positive selection have been found in African American and Puerto Rican populations^39^. Since the mtDNA of admixed Latino populations is predominantly of Native American origin, and their nuclear genome reflects a gradient of Native American, European and African components, admixed Latinos, such as our group of patients, would present a great opportunity to study the impact MND on the 22q11DS phenotypes in the future.

## Data Availability

Raw sequencing data is available under Bio Project accession number PRJNA636010. Reviewer link: https://dataview.ncbi.nlm.nih.gov/object/PRJNA636010?reviewer=r8tqn3odmkvdh9cdhhb3ujgdj0

## ACKNOWLEDGMENTS

We would like to thank all participating patients, without whom this study would not have been possible. B.R-J. would like to thank Analia Cuiza and Gonzalo Encina, for their technical support in patient enrolment and sample processing, respectively. This work was funded by the National Agency for Research and Development (ANID) FONDECYT grant 3170280 and 1171014.

## AUTHOR CONTRIBUTIONS

B.R.-J. and G.M.R. conceived the project. B.R.-J analyzed the data and drafted the manuscript. M.G.O., V.H., and A.G. enrolled patients, performed clinical phenotyping and follow up, and processed samples. M.G.O. and G.M.R. collaborated in manuscript writing. All authors reviewed and accepted the final version.

## COMPETING INTERESTS

The authors declare no competing interests.

## Supplementary material

**Supplementary figure 1.**
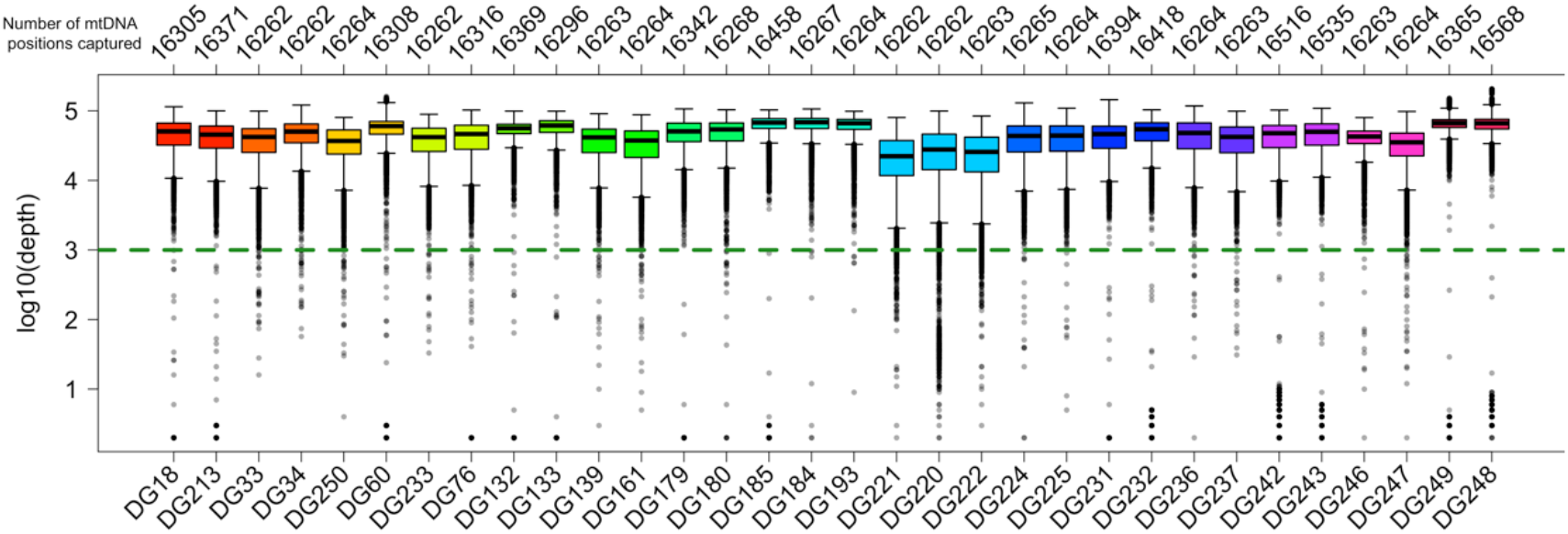
Per sample mtDNA sequencing depth distribution. Samples are colored according to familial relationship.

**Supplementary figure 2.**
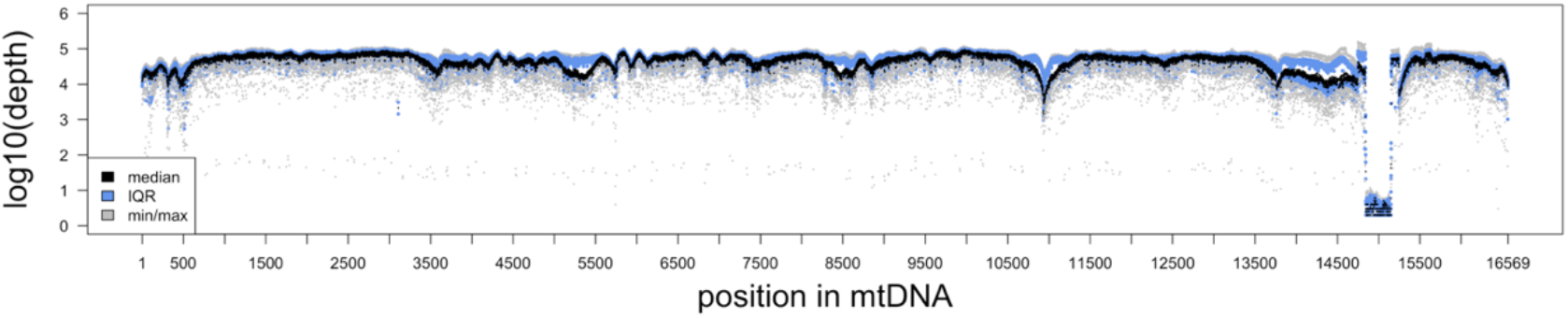
Per mtDNA position depth distribution. Each position in the human mtDNA (16,569 positions) is represented by a boxplot according to the colors indicated in the legend.

**Supplementary figure 3.**
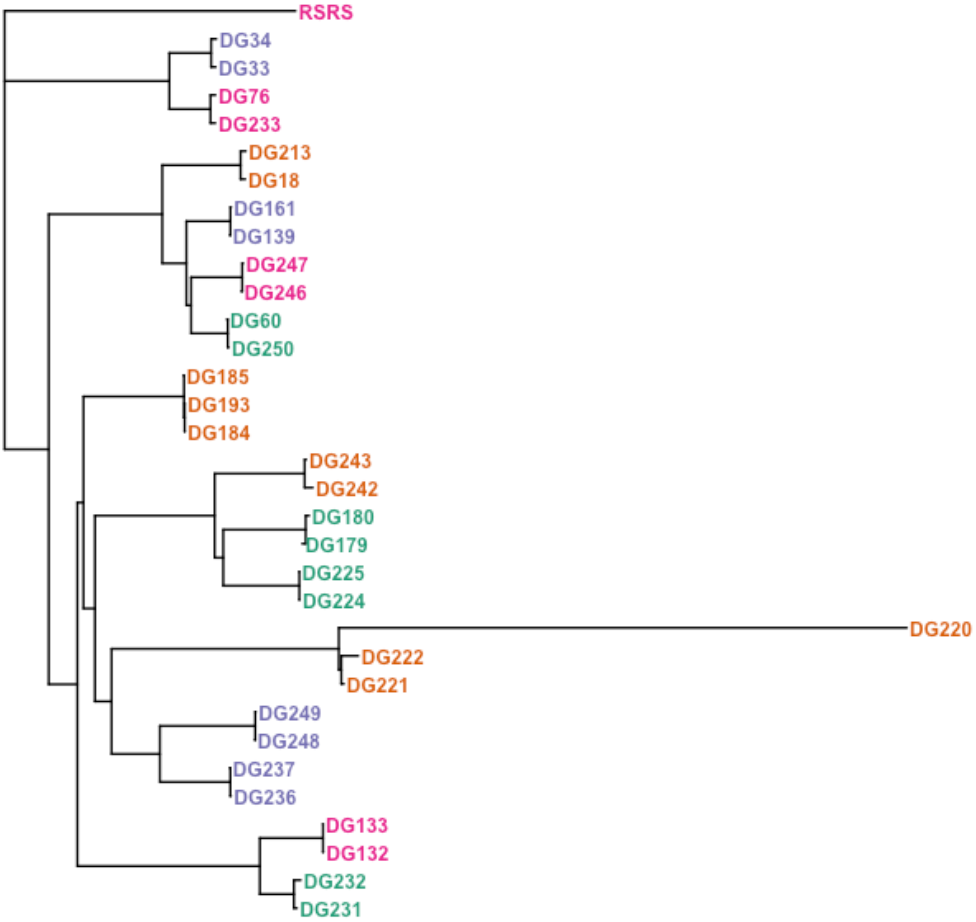
Pairwise distance neighbor joining tree reflects maternal relationships. The tree has been rooted with the Reconstructed Sapiens Reference Sequence (RSRS)^40^.

**Supplementary figure 4.**
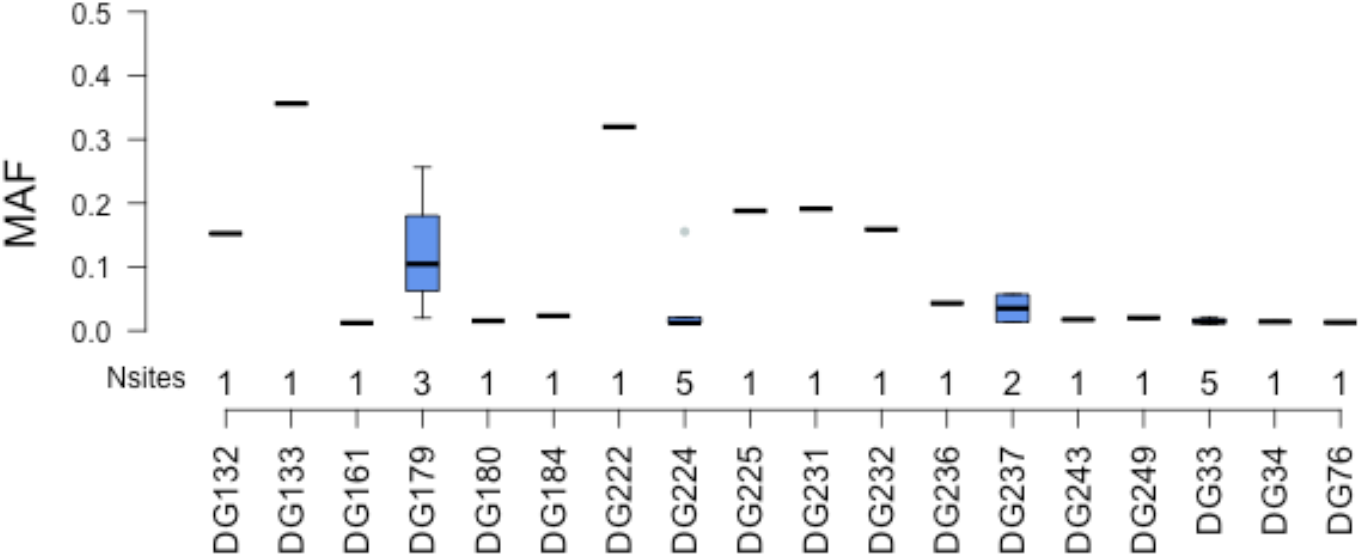
Minor allele frequency distribution of samples with heteroplasmic sites. MAF: Minor allele frequency.

**Supplementary figure 5.**
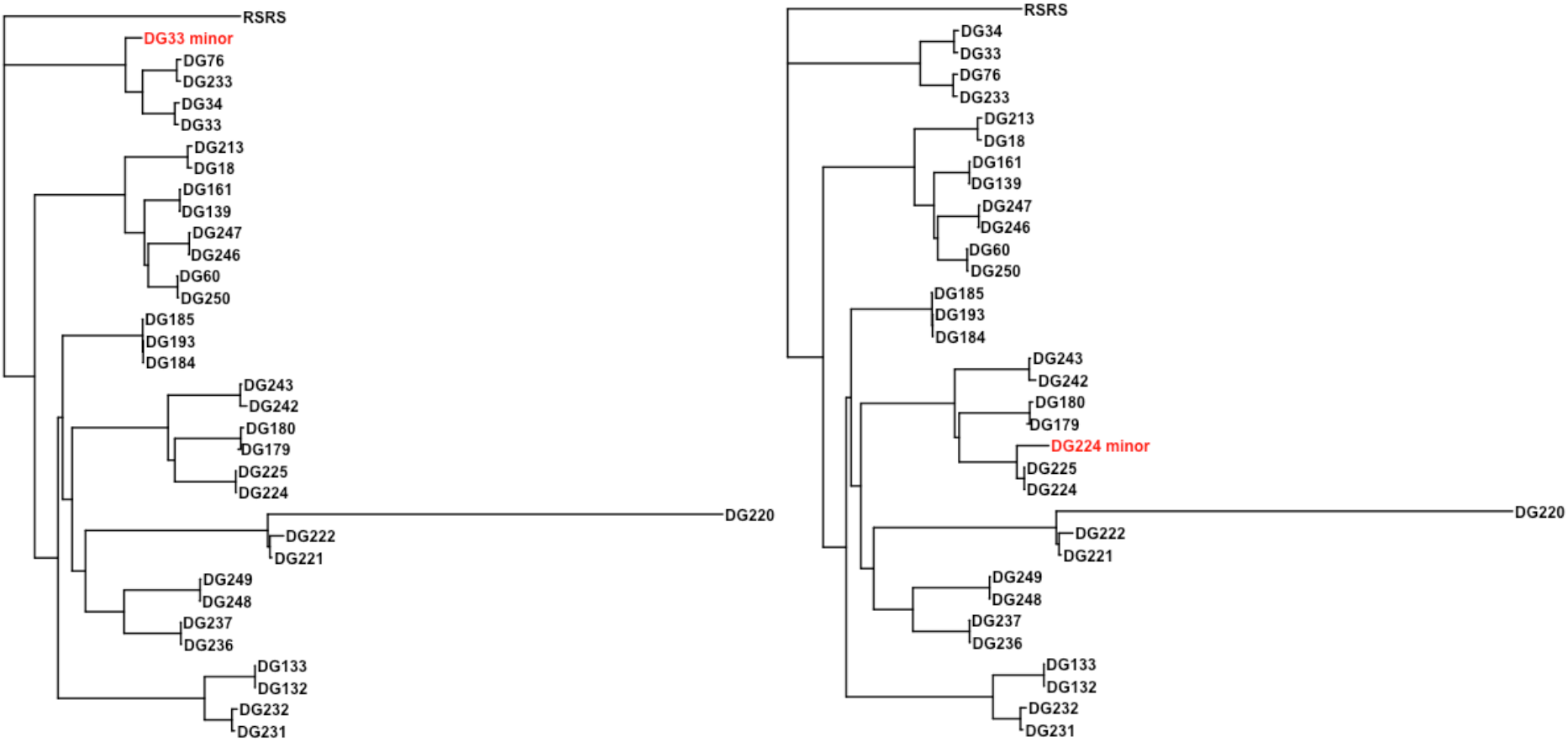
Minor allele sequences cluster with expected related sequences. Samples DG33 and DG224, both had 5 heteroplasmic sites each with apparently non-random minor allele frequencies (the five sites had very similar AF), but they do not cluster with unrelated sequences.

**Supplementary table 1.** mtDNA haplogroups. We confirmed maternal relationships between mother-child pairs based on their haplogroup. Conf: posterior probability of haplogroup assignment.

| Pair | Mother | Child | Hap_mother | Conf_mother | Hap_child | Conf_child |
| --- | --- | --- | --- | --- | --- | --- |
| 1 | DG213 | DG18 | A2+(64)+@153 | 0.87 | A2+(64)+@153 | 0.88 |
| 2 | DG34 | DG33 | D1j1a | 0.94 | D1j1a | 0.93 |
| 3 | DG60 | DG250 | A2+(64) | 0.97 | A2+(64) | 0.97 |
| 4 | DG76 | DG233 | D1j1a1 | 0.98 | D1j1a1 | 0.97 |
| 5 | DG133 | DG132 | T2b3+151 | 0.93 | T2b3+151 | 0.93 |
| 6 | DG161 | DG139 | A2+(64) | 0.93 | A2+(64) | 0.93 |
| 7 | DG180 | DG179 | B2 | 0.80 | B2 | 0.80 |
| 8 | DG184 | DG185 | H3ao1 | 0.93 | H3ao1 | 0.93 |
| 9 | DG184 | DG193 | H3ao1 | 0.93 | H3ao1 | 0.93 |
| 10 | DG220 | DG221 | U1a1a3 | 0.69 | U1a1a3 | 0.90 |
| 11 | DG220 | DG222 | U1a1a3 | 0.69 | U1a1a3 | 0.89 |
| 12 | DG225 | DG224 | B2 | 0.78 | B2 | 0.78 |
| 13 | DG232 | DG231 | T2b | 0.95 | T2b | 0.96 |
| 14 | DG237 | DG236 | U5b1+16189+@16192 | 0.96 | U5b1+16189+@16192 | 0.96 |
| 15 | DG243 | DG242 | B2b3a | 0.91 | B2b3a | 0.91 |
| 16 | DG247 | DG246 | A2+(64) | 0.93 | A2+(64) | 0.93 |
| 17 | DG248 | DG249 | U5a1a1 | 0.96 | U5a1a1 | 0.96 |

**Supplementary table 2.** Allele frequency change (∆AF) between a mother and her child for all heteroplasmic sites identified. mid: mother id. cid: child id. pos: position. nt: allele. mc: mother allele count. md: mother site depth. AFm: Allele frequency mother. cc: child allele count. cd: child site depth. AFc: Allele frequency child. ∆AF: Delta allele frequency (child – mother). ht: heart phenotype. pl: palate phenotype.

| mid | cid | pos | nt | mc | md | AFm | cc | cd | AFc | ΔAF | ht | pl |
| --- | --- | --- | --- | --- | --- | --- | --- | --- | --- | --- | --- | --- |
| DG34 | DG33 | 6802 | G | 529 | 37167 | 1,E-02 | 1 | 43112 | 2,32E-05 | -1,42E-02 | con | con |
| DG34 | DG33 | 10873 | T | 7 | 9519 | 7,E-04 | 170 | 11859 | 1,43E-02 | 1,36E-02 | con | con |
| DG34 | DG33 | 14668 | C | 15 | 34435 | 4,E-04 | 54 | 4713 | 1,15E-02 | 1,10E-02 | con | con |
| DG34 | DG33 | 14783 | T | 11 | 23194 | 5,E-04 | 143 | 13425 | 1,07E-02 | 1,02E-02 | con | con |
| DG34 | DG33 | 15301 | G | 1 | 50276 | 2,E-05 | 194 | 10341 | 1,88E-02 | 1,87E-02 | con | con |
| DG34 | DG33 | 15326 | G | 2 | 47401 | 4,E-05 | 307 | 14117 | 2,17E-02 | 2,17E-02 | con | con |
| DG76 | DG233 | 15786 | C | 867 | 66261 | 1,E-02 | 0 | 62238 | 0,00E+00 | -1,31E-02 | dis | dis |
| DG133 | DG132 | 9507 | C | 21847 | 61348 | 4,E-01 | 8407 | 55188 | 1,52E-01 | -2,04E-01 | dis | dis |
| DG161 | DG139 | 5492 | C | 362 | 29462 | 1,E-02 | 5 | 30038 | 1,66E-04 | -1,21E-02 | dis | dis |
| DG180 | DG179 | 150 | T | 298 | 19275 | 2,E-02 | 4900 | 19122 | 2,56E-01 | 2,41E-01 | dis | con |
| DG180 | DG179 | 1316 | C | 2 | 24450 | 8,E-05 | 2255 | 21577 | 1,05E-01 | 1,04E-01 | dis | con |
| DG180 | DG179 | 5054 | A | 37 | 68532 | 5,E-04 | 1410 | 70310 | 2,01E-02 | 1,95E-02 | dis | con |
| DG184 | DG185 | 15591 | A | 630 | 26908 | 2,E-02 | 20 | 39996 | 5,00E-04 | -2,29E-02 | dis | dis |
| DG184 | DG193 | 15591 | A | 630 | 26908 | 2,E-02 | 19 | 32685 | 5,81E-04 | -2,28E-02 | dis | dis |
| DG220 | DG221 | 16290 | C | 6712 | 7398 | 9,E-01 | 6713 | 6716 | 1,00E+00 | 9,23E-02 | con | dis |
| DG220 | DG222 | 16290 | C | 6712 | 7398 | 9,E-01 | 4390 | 13735 | 3,20E-01 | -5,88E-01 | con | dis |
| DG225 | DG224 | 6190 | A | 117 | 17184 | 7,E-03 | 94 | 8443 | 1,11E-02 | 4,32E-03 | con | con |
| DG225 | DG224 | 10075 | C | 0 | 80165 | 0,E+00 | 1636 | 77194 | 2,12E-02 | 2,12E-02 | con | con |
| DG225 | DG224 | 12315 | A | 10 | 46357 | 2,E-04 | 509 | 44701 | 1,14E-02 | 1,12E-02 | con | con |
| DG225 | DG224 | 12457 | A | 25 | 7323 | 3,E-03 | 34 | 2796 | 1,22E-02 | 8,75E-03 | con | con |
| DG225 | DG224 | 16182 | A | 432 | 2299 | 2,E-01 | 423 | 2721 | 1,55E-01 | -3,25E-02 | con | con |
| DG232 | DG231 | 4136 | G | 6158 | 38779 | 2,E-01 | 6499 | 34006 | 1,91E-01 | 3,23E-02 | dis | con |
| DG237 | DG236 | 5755 | G | 75 | 1316 | 6,E-02 | 76 | 1764 | 4,31E-02 | -1,39E-02 | con | con |
| DG237 | DG236 | 15355 | A | 366 | 27184 | 1,E-02 | 6 | 30354 | 1,98E-04 | -1,33E-02 | con | con |
| DG243 | DG242 | 15431 | A | 689 | 38994 | 2,E-02 | 15 | 36189 | 4,14E-04 | -1,73E-02 | dis | con |
| DG248 | DG249 | 9941 | G | 6 | 85309 | 7,E-05 | 1615 | 79897 | 2,02E-02 | 2,01E-02 | dis | dis |

## Notes

### Competing Interest Statement

The authors have declared no competing interest.

### Author Declarations

Comite Etico Cientifico, Facultad de Medicina, Universidad del Desarrollo, Chile. Ref. 2017-56 Comite Hospitalario de etica, Hospital Garrahan, Argentina. Ref. 1127

